# Clinical validation of the quantitative Siemens SARS-CoV-2 spike IgG assay (sCOVG) reveals improved sensitivity and a good correlation with virus neutralization titers

**DOI:** 10.1101/2021.02.17.21251907

**Authors:** Christian Irsara, Alexander E. Egger, Wolfgang Prokop, Manfred Nairz, Lorin Loacker, Sabina Sahanic, Alex Pizzini, Thomas Sonnweber, Barbara Holzer, Wolfgang Mayer, Harald Schennach, Judith Loeffler-Ragg, Rosa Bellmann-Weiler, Boris Hartmann, Ivan Tancevski, Günter Weiss, Christoph J. Binder, Markus Anliker, Andrea Griesmacher, Gregor Hoermann

## Abstract

**Objectives:** Severe Acute Respiratory Syndrome Coronavirus 2 (SARS-CoV-2) infections cause Coronavirus Disease 2019 (COVID-19) and induce a specific antibody response. Serological assays detecting IgG against the receptor binding domain (RBD) of the spike (S) protein are useful to monitor the immune response after infection or vaccination. The objective of our study was to evaluate the clinical performance of the Siemens SARS-CoV-2 IgG (sCOVG) assay.

**Methods:** Sensitivity and specificity of the Siemens sCOVG test were evaluated on 178 patients with SARS-CoV-2-infection and 160 pre-pandemic samples in comparison with its predecessor test COV2G. Furthermore, correlation with virus neutralization titers was investigated on 134 samples of convalescent COVID-19 patients.

**Results:** Specificity of the sCOVG test was 99.4% and sensitivity was 90.5% (COV2G assay 78.7%; p<0.0001). S1-RBD antibody levels showed a good correlation with virus neutralization titers (r=0.843; p<0.0001) and an overall qualitative agreement of 98.5%. Finally, median S1-RBD IgG levels increase with age and were significantly higher in hospitalized COVID-19 patients (median levels general ward: 25.7 U/ml; intensive care: 59.5 U/ml) than in outpatients (3.8 U/ml; p<0.0001).

**Conclusions:** Performance characteristics of the sCOVG assay have been improved compared to the predecessor test COV2G. Quantitative SARS-CoV-2 S1-RBD IgG levels could be used as a surrogate for virus neutralization capacity. Further harmonization of antibody quantification might assist to monitor the humoral immune response after COVID-19 disease or vaccination.

## Introduction

Coronavirus disease 2019 (COVID-19), which is caused by the Severe Acute Respiratory Syndrome Coronavirus 2 (SARS-CoV-2) [1, 2] was declared pandemic by the WHO on March 11, 2020 [3] and is still challenging the health systems and governments all over the world. As the development of vaccines evolves very rapidly [4] and vaccines are continually approved [5-8] there is a growing need for highly specific and sensitive serologic assays not only for supporting COVID-19 diagnosis in the individual patient and for seroprevalence studies but also to estimate the quality and quantity of humoral immune response to vaccination.

Serologic SARS-CoV-2 tests can be categorized by the assay type (neutralization assays [9] vs. immunoassays [10]), the antibody isotype (IgA, IgG, IgM or total antibodies [11]), and type of viral antigen detected (Nucleocapsid[N]-[12], Spike[S]-protein[13], Receptor Binding Domain [RBD] [14] of the S-protein) and the type of result reporting (qualitative, semi-quantitative, quantitative). The quantification of the humoral immune response to SARS-CoV-2 virus infection or vaccination in large patient cohorts should be performed by an immunoassay, which shows a good correlation to a neutralization assay [15-17]. However, the international harmonization of SARS-CoV-2 serologic assays regarding quantitative values, especially in vaccine recipients, is still pending. The first step toward that goal is the establishment of the first WHO International Standard and Reference Panel for anti-SARS-CoV-2 antibody [18].

Recently, we reported a clinical evaluation of the Siemens SARS-CoV-2 IgG (COV2G) in comparison to three other fully automated SARS-CoV-2 chemiluminescence immunoassays on high throughput random access analysers (Roche Elecsys Anti-SARS-CoV-2, Abbott SARS-CoV-2 IgG, Siemens SARS-CoV-2 total). In that study, the sensitivity of the Siemens COV2G test (78.8%) was unexpectedly low and inferior to that of the other assays (range 90.8% to 93%) [19]. In the meantime, Siemens has launched a new SARS-CoV-2 IgG test (sCOVG) by November 18, 2020. This newly filed assay also detects the S1-RBD antigen and is intended to be used for qualitative and quantitative detection of SARS-CoV-2 IgG including neutralizing antibodies [20]. In this study, we aimed to clinically validate this new Siemens sCOVG assay with a particular emphasis on sensitivity using the same samples as in our prior study to ascertain maximal comparability with the previous Siemens COV2G assay. In consideration of the ongoing vaccination programs, we also focussed on validating the potential of the sCOVG assay to quantify IgG antibodies and its correlation with a neutralization assay.

## Materials and methods

### Patients and study design

The present study was performed at the University Hospital of Innsbruck as part of the clinical evaluation of different SARS-CoV-2 serologic assays. All procedures performed in the present study involving human participants were in accordance with the ethical standards of the Institutional and/or National Research Committee and with the 1964 Helsinki declaration and its later amendments and were approved by the ethics committee of the Medical University of Innsbruck (ethics commission numbers: 1103/2020, 1167/2020).

193 patients with reverse transcription polymerase chain reaction (RT-PCR)-confirmed SARS-CoV-2 infection dating between March and August, 2020, were screened for this study. All samples have been previously tested with the Siemens SARS-CoV-2 IgG assay (COV2G) [19]. 15 patients (7.8%) were excluded, as no sample material was available for analysis. The patients’ characteristics of the remaining 178 patients are shown in Table 1. Sensitivity in the investigated cohort was evaluated using one sample per patient dating ≥14 days after disease onset and the sample closest to day 28 after disease onset was chosen. Disease onset was defined as onset of clinical symptoms compatible with COVID-19 infection (n=156, 88%), or as the first positive SARS-CoV-2 RT-PCR when symptom onset was not available (n=22, 12%). Furthermore, 134 samples of RT-PCR-confirmed COVID-19 patients (only one sample per patient) from the CovILD-study cohor t[21] were tested in comparison to a SARS-CoV-2 neutralization assay. The patients’ characteristics of this cohort are shown in Supplemental Table S1). Of those 134 samples from different patients, 52 (39%) overlapped with the 178 samples of the sensitivity analysis described above. In addition, 160 pre-pandemic samples were used to verify specificity. Finally, an intravenous immunoglobulin formulation (Privigen®, 100 mg/ml, CSL Behring AG, Bern, Switzerland) composed of pre-pandemic pooled immunoglobulins (mainly IgG) of a large number of healthy donors from the US, which should by definition yield negative SARS CoV-2 antibody results, was tested for possible false positive cross reactions.

**Table 1.** Characteristics of COVID-19 patients (sensitivity analysis cohort)

|  |  | Total |  | Outpatient |  | Hospitalized<br>General ward |  | Hospitalized<br>Intensive care |  |
| --- | --- | --- | --- | --- | --- | --- | --- | --- | --- |
|  |  | Absolute | % | Absolute | % | Absolute | % | Absolute | % |
| **Number** | n | 178 | 100% | 65 | 37% | 85 | 48% | 28 | 16% |
| **Age (years)** | min | 20 |  | 20 |  | 27 |  | 44 |  |
|  | max | 95 |  | 83 |  | 95 |  | 79 |  |
|  | median (IQR) | 56 (43- 68) |  | 39 (28- 56) |  | 63 (54- 76) |  | 59 (54- 65) |  |
| **Sex** | female | 65 | 37% | 26 | 40% | 32 | 38% | 7 | 25% |
|  | male | 113 | 63% | 39 | 60% | 53 | 62% | 21 | 75% |
| **Symptom onset known** | yes | 156 | 88% | 47 | 72% | 82 | 96% | 27 | 96% |
|  | no | 22 | 12% | 18 | 28% | 3 | 4% | 1 | 4% |
| **Time between symptom onset and PCR (days)<sup>a</sup>** | median (IQR) | 5 (2-8) |  | 2 (1-6) |  | 6 (3-8) |  | 5 (2-7) |  |
| **Time between disease onset and blood draw (days)<sup>b</sup>** | median (IQR) | 47 (24-61) |  | 58 (47-68) |  | 28 (19-59) |  | 28 (25-31) |  |

<sup>a</sup> Refers to the first positive SARS-CoV-2 PCR of the patient; <sup>b</sup> refers to the representative sample used in the sensitivity analysis. Abbreviations: IQR, interquartile range, PCR, polymerase chain reaction.

### Siemens SARS-CoV-2 IgG (sCOVG) assay

Blood samples were prepared as described previously [19]. We evaluated the Siemens SARS-CoV-2 IgG assay (sCOVG) on the Siemens ADVIA Centaur XP platform (Siemens, Munich, Germany). All samples were processed according to the manufacturer’s procedures with the specified controls and calibrators by trained laboratory staff. Results of SARS-CoV-2 IgG are given as U/ml, whereby the cut-off for positivity is defined as ≥ 1.0 U/ml. The manufacturer reports a range of quantification of 0.5-150.0 U/ml, which may be extended to 750.0 U/ml upon automated 1:5 predilution with the diluent provided by the company. In the sensitivity analysis cohort (n=178) no predilution was necessary, while in the neutralization assay cohort (n=134) six samples had to be diluted as described. Additional test characteristics given in the manufacturer’s product information are summarized in Supplemental Table S2. Precision was evaluated by repeatedly measuring the positive control of the assay.

### SARS-CoV-2 neutralization assay

Neutralizing antibody titers in human serum and plasma were determined using a tissue culture infectious dose (TCID_50_) assay for authentic SARS-CoV-2 virus on Vero 76 clone E6 cells as described [22]. A titer of at least 1:4 defined a positive result in the assay.

### Data analysis and statistics

Statistical analyses were performed using MedCalc, version 19.6.1 (MedCalc Ltd., Ostend, Belgium) and Excel 2016 (Microsoft, Redmont, USA). 95% confidence intervals (CI) for proportions were calculated according to the Clopper-Pearson exact method. The difference between categorical data was assessed using Chi-square test (McNemar’s test for paired data, “N-1” Chi-squared test for unpaired proportions). The difference between quantitative data was assessed using Mann-Whitney test for two groups and the Kruskal-Wallis test followed by Dunn’s post-hoc test for more than two groups. The correlation of quantitative antibody results and the neutralization assay titers was assessed using Spearman’s coefficient of rank correlation (rho). Statistical significance was defined at a level of 0.05.

## Results

### Precision

Intra-assay and inter-run precision for the Siemens SARS-CoV-2 IgG (sCOVG) were 3.8% (n=10) and 6.1% (n=10), respectively.

### Specificity

Out of 160 pre-pandemic samples, one tested borderline positive in the Siemens sCOVG assay (Index: 1.24), resulting in a specificity of 99.4% (95% CI: 96.6-100.0%, Table 2), which is in line with the manufacturer’s claims (99.90%, 95% CI: 99.64-99.99%). The measurement of the undiluted pre-pandemic intravenous immunoglobulin formulation Privigen® yielded a borderline positive result (1.28 U/ml). However, in a more physiologic dilution of 1:50, it yielded a clearly negative result (Supplemental Table S3).

**Table 2.**
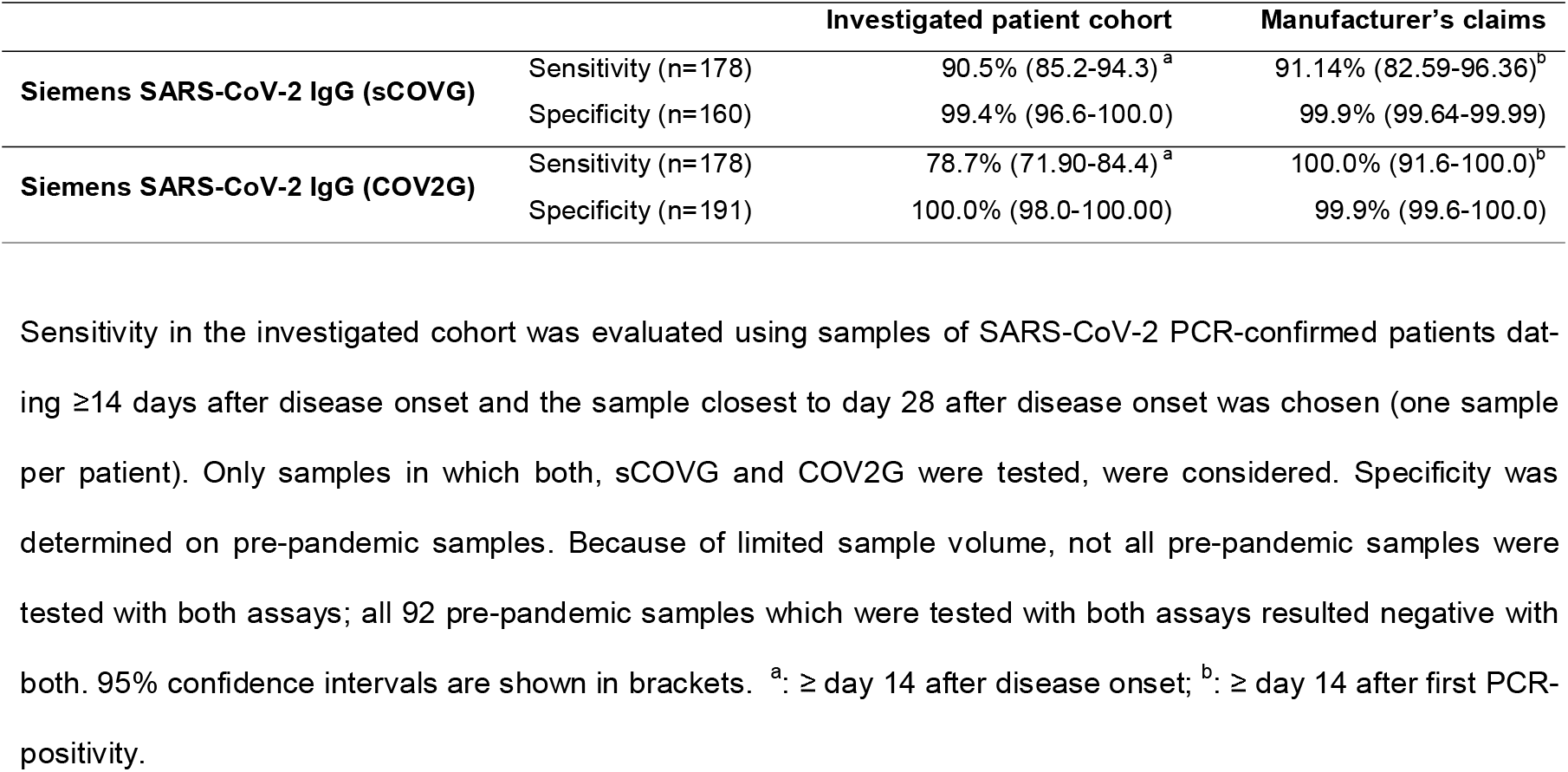
Sensitivity and specificity of the Siemens sCOVG and COV2G assays

### Sensitivity

Out of 178 patients with RT-PCR confirmed SARS-CoV-2 infection, 161 tested positive and 17 negative. Thus the sensitivity was 90.5% (95% CI: 85.2-94.3%) for the sCOVG assay (Table 2, Figure 1A). When we compared these results with those of the previous COV2G assay of the same samples, we found the new sCOVG assay was significantly more sensitive (p<0.0001; Table 2, Figure 1A, Supplemental Figure S1).

**Figure 1.**
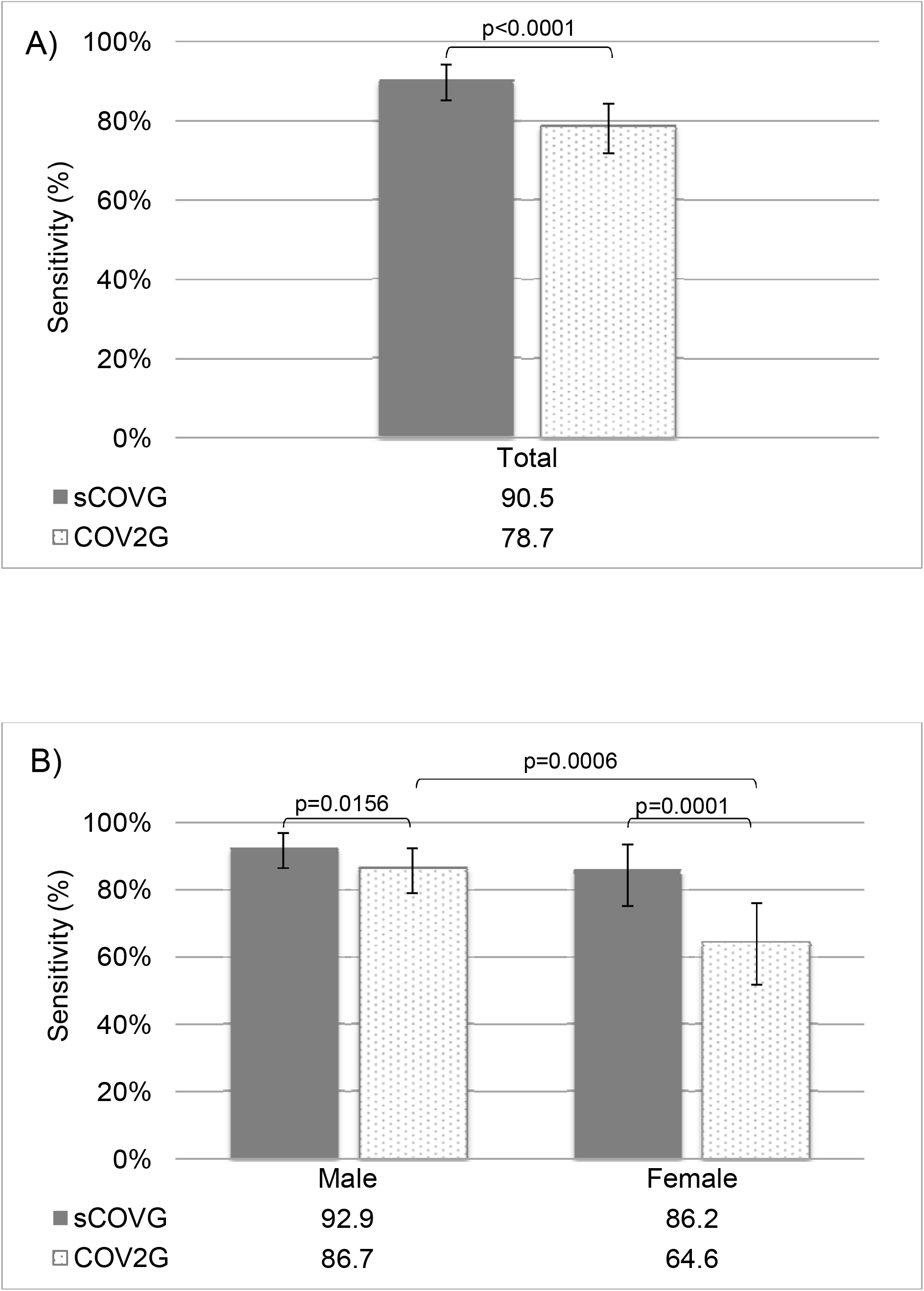
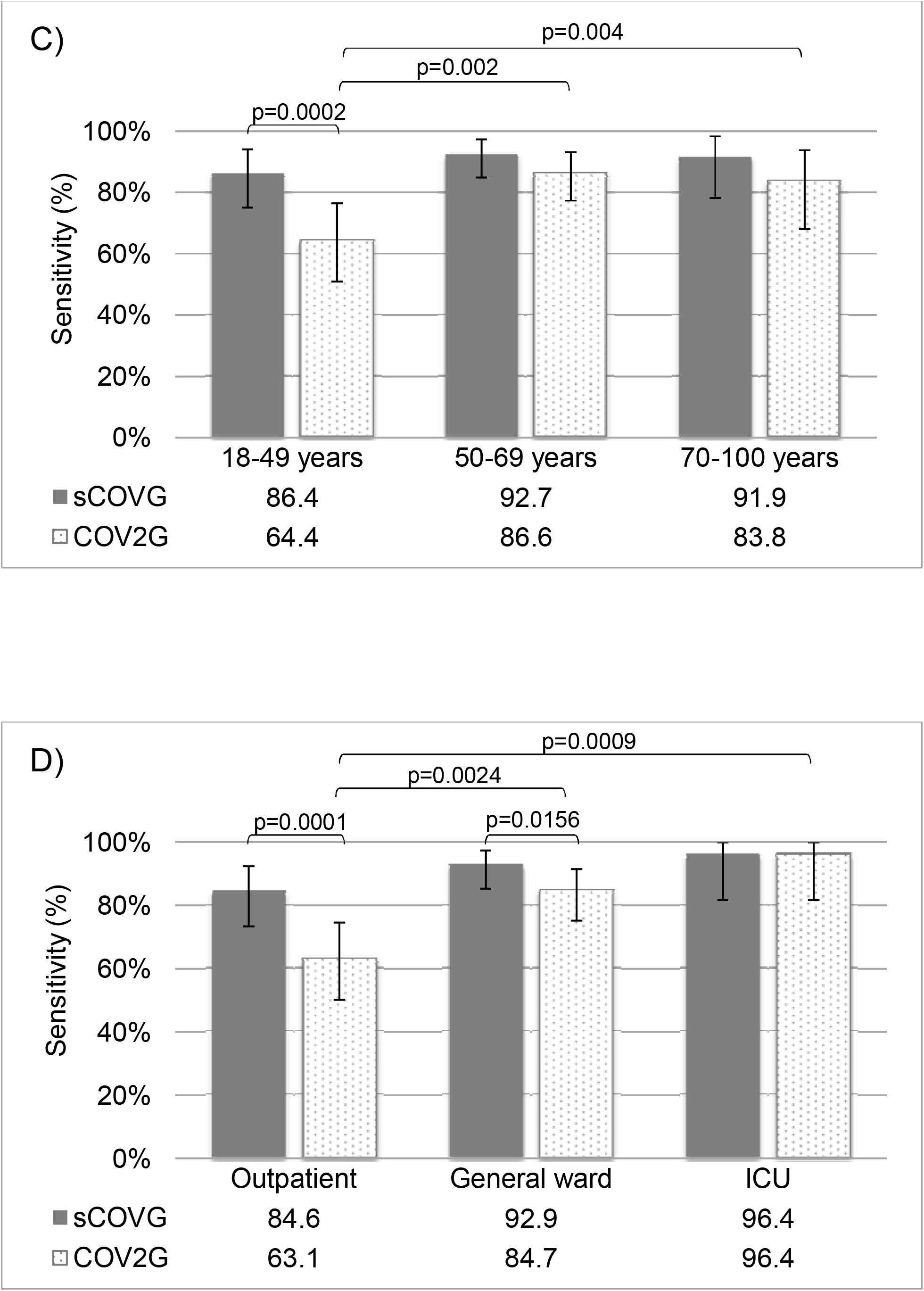
Sensitivity, including subgroup analyses according to gender, age and severity of disease. Comparison of the sensitivity of the two investigated assays (Siemens SARS-CoV-2 IgG sCOVG in grey, and COV2G dotted) in representative samples of 178 patients with PCR-confirmed COVID-19 (A). Additionally, results were analysed stratified for gender (B), age (C) and severity of disease (D). The sample numbers for the different cohorts were for female n=65, male n=113, age 18-49 years n=59, age 50-69 years n=82, age 70-100 years n=37, outpatient n=65, patients at the general ward n=85 and patients requiring intensive care n=28.

When stratifying for gender, age and severity of disease (Figure 1B-D, Supplemental Table S4), the sCOVG assay also had a significantly higher sensitivity than the COV2G assay in male (92.9% vs. 86.7%, p=0.0156), female (86.2% vs. 64.6%, p=0.0001), patients aged 18-49 years (86.4% vs. 64.4%, p=0.0002), outpatients (84.6% vs. 63.1%, p=0.0001) and patients at the general ward (92.9% vs. 84.7%, p=0.0156).

### Correlation to the previous Siemens COV2G assay

When comparing the quantitative index value raw data of the 178 sensitivity samples, the Spearman’s coefficient of rank correlation (r) was 0.919 (95%CI: 0.892-0.939, p<0.0001) between the sCOVG and the COV2G assay. However, the scatter diagram (Supplemental Figure S2) shows that in 21/178 (11.8%) samples, the manufacturer’s cut-off index (COI) for positivity was exceeded only in the sCOVG but not the COV2G (lower right quadrant), while vice versa no sample exceeded the COI for positivity in the COV2G but not in the sCOVG assay. 17/178 (9.6%) samples showed index values below the COI (lower left quadrant) and 140/178 (78.7%) samples showed an index value above the COI (upper right quadrant) with both assays, respectively. In summary, the raw data index of the formerly COV2G assay correlates with the quantitative results of the new sCOVG test but the new test is better in discriminating values in samples with higher antibody concentrations.

### Correlation with SARS-CoV-2 neutralization titers

Samples of 134 patients of the CovILD-study cohort (Supplemental Table S1) were tested for virus neutralization capacity using a TCID_50_ assay for authentic SARS-CoV-2 virus. 126 patients (94%) tested positive for the presence of virus neutralizing antibodies. The sCOVG assay showed an overall qualitative agreement of 98.5% to the results of the SARS-CoV-2 neutralization test (Supplemental Table S5). Only two samples (1.5%) yielded discordant results being positive in the neutralization assay but not in the sCOVG test with 0.3 IU/ml and 0.6 IU/ml. Importantly, also the quantitative values of antibodies against the RBD of the S-Protein of SARS-CoV-2 showed a good correlation to virus neutralization titers (Spearman’s r=0.843, 95%CI: 0.785-0.885, p<0.0001; Figure 2).

**Figure 2.**
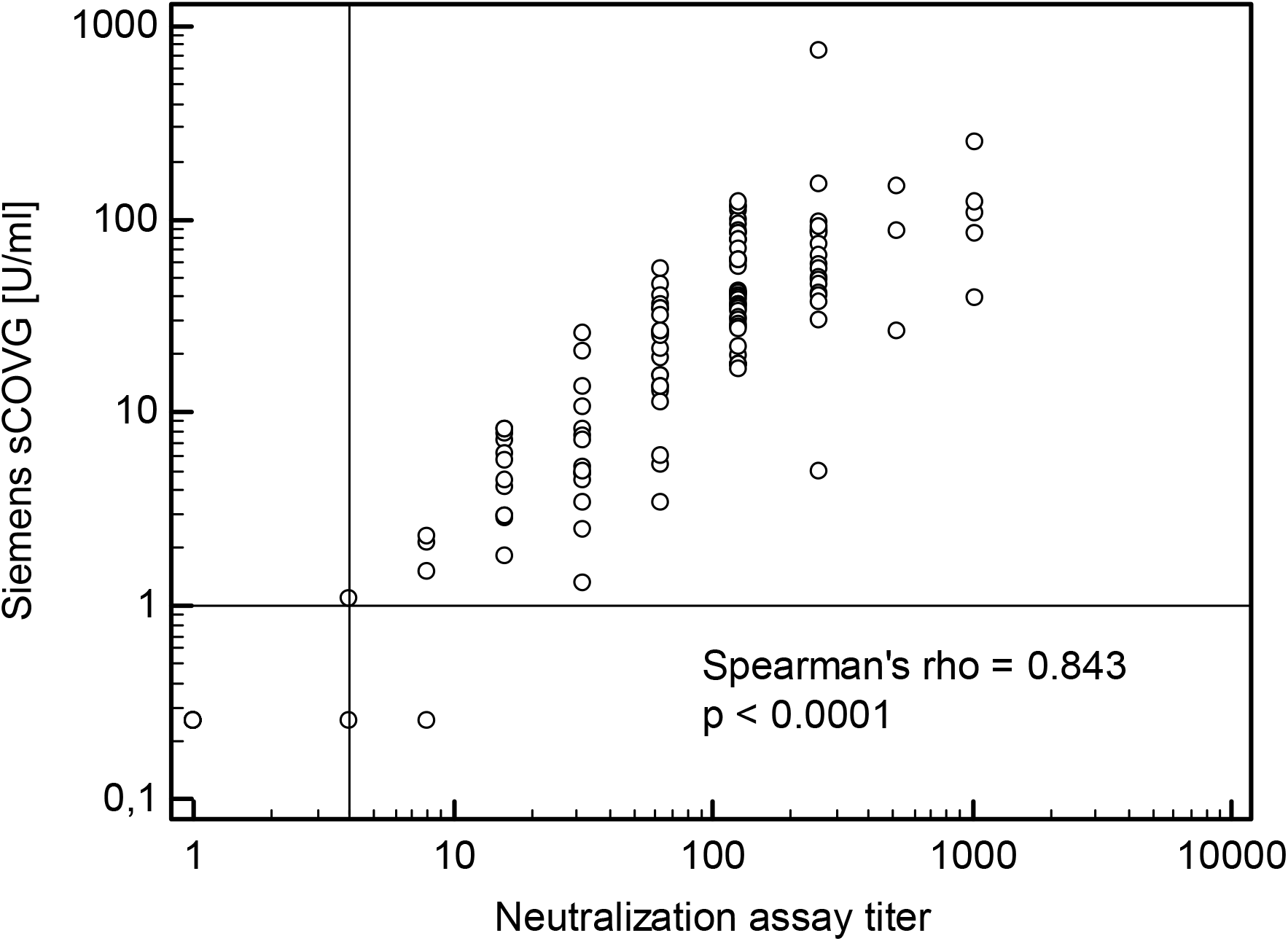
Correlation between the quantitative Siemens SARS-CoV-2 IgG (sCOVG, U/ml) value and the neutralization assay titer in 134 samples of different SARS-CoV-2 RT-PCR confirmed patients. The horizontal line shows the COI for positivity of the sCOVG assay (≥1.0 U/ml), the vertical line shows the cut off for positivity of the neutralization assay (titer level ≥ 4). For this illustration, values below the limit of quantification (LoQ, 0.5) in the sCOVG test were set as 0.25 U/ml, neutralization assay titers < 4 were set as 1 and titers > 512 as 1,024.

### Patients with severe COVID-19 show higher SARS-CoV-2 S-RBD IgG levels

Finally, we used the 178 samples of the initial sensitivity cohort in an exploratory analysis to study the impact of patients’ and disease characteristics on the quantitative level of IgG antibodies against the S-RBD of SARS-CoV-2 as determined by the Siemens sCOVG assay. Female subjects showed significantly lower quantitative antibody values (n=65, median 5.59 U/ml, IQR 1.84-29.73 U/ml) as compared to males (n=113, median 23.88 U/ml, IQR 5.64-48.79 U/ml; p=0.0003; Figure 3A). Surprisingly, patients aged 18-49 years also showed markedly lower values (n=59, median 4.1 U/ml, IQR 3.2-8.0 U/ml) than patients aged 50-69 years (n=82, median 27.2 U/ml, IQR 7.7-53.1 U/ml, p<0.0001) or patients aged 70-100 years (n=37, median 25.7 U/ml, IQR 4.5-46.3 U/ml, P=0.0032). The difference between patients aged 50-69 years and patients aged 70-100 years was not significant (Figure 3B).

**Figure 3.**
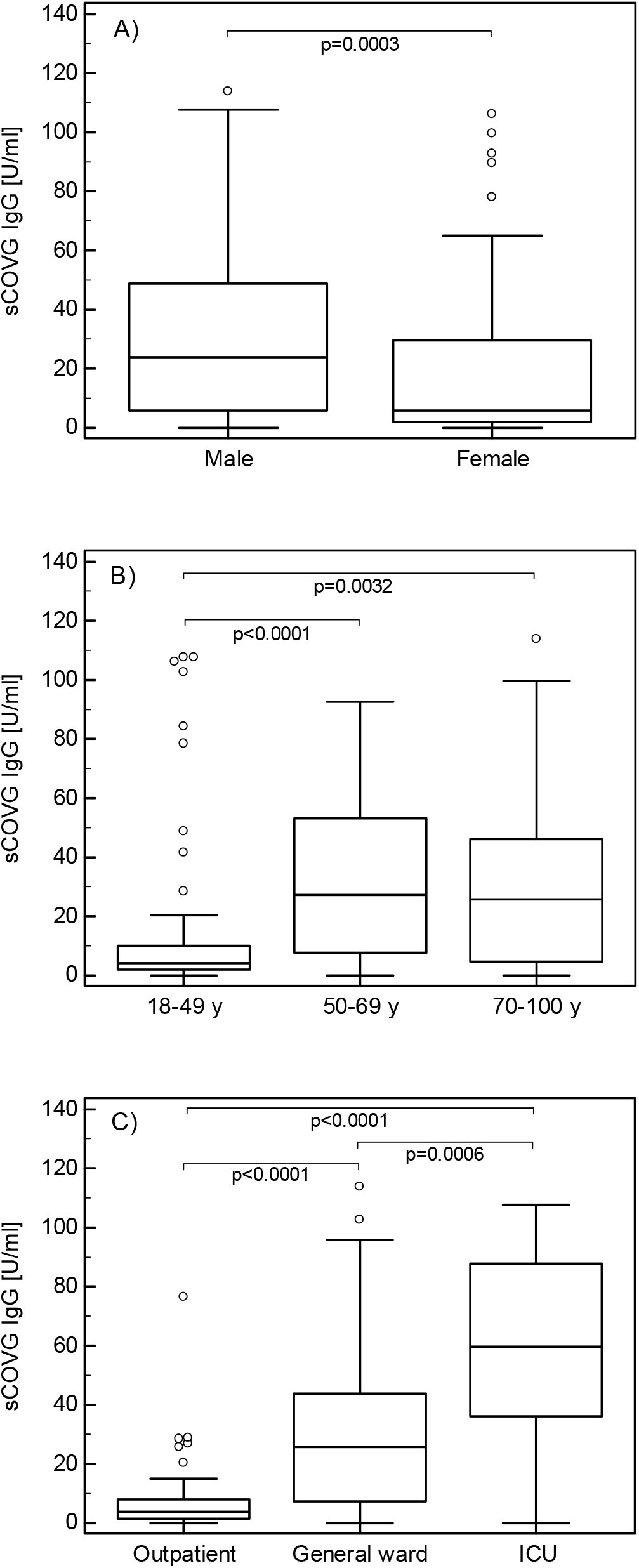
A-C. Comparison of the quantitative values (U/ml) of the Siemens SARS-CoV-2 IgG (sCOVG) assay depending on gender, age and severity of disease. The quantitative values of the 178 samples of different SARS-CoV-2 RT-PCR-confirmed patients were analysed according to gender (A), age (B) and severity of disease (C). The sample numbers for the different cohorts were for female n=65, male n=113, age 18-49 years n=59, age 50-69 years n=82, age 70-100 years n=37, outpatient n=65, patients at the general ward n=85 and patients requiring intensive care (ICU) n=28.

We then asked whether this age dependent increase in serum antibody levels could be linked to disease severity, which is more prevalent in elderly subjects [21, 23]. Outpatients showed clearly and significantly lower antibody values (n=65, median 3.8 U/ml, IQR 1.4-8.2 U/ml) than hospitalized patients either at the general ward (n=85, median 25.7 U/ml, IQR 7.5-43.9 U/ml, p<0.0001) or at the ICU (n=28, median 59.5 U/ml, IQR 36.0-87.8 U/ml, p<0.0001). Also, the difference between the patients at the general ward and ICU was statistically significant (p=0.0006; Figure 3C). As the time between the onset of disease and the blood draw for antibody testing was not equal for all cohorts (Supplemental Figure S3) and the kinetics of humoral immune response might influence our results, we restricted the analysis to samples which were drawn at least 30 days after disease onset (n=112). When considering only those samples, still a similar picture was seen: outpatients (n=62, median 3.5 U/ml, IQR 1.3-7.9 U/ml) showed significantly lower antibody values than patients who were previously treated at the general ward (n=40, median 17.6 U/ml, IQR 5.8-39.4 U/ml, p<0.0001) or at the ICU (n=10, median 57.9 U/ml, IQR 23.9-89.6 U/ml, p<0.0001). In those samples dated ≥ 30 days after symptom onset, patients aged 18-49 years (n=51, median 3.6 U/ml, IQR 1.8-8.7 U/ml) showed significantly lower antibody values than patients aged 50-69 years (n=47, median 12.6 U/ml, IQR 5.2-34.1 U/ml, p=0.0019).

## Discussion

Our clinical evaluation of the Siemens SARS-CoV-2 IgG (sCOVG) showed improved sensitivity compared to the previous COV2G test. Quantitative results for S-RBD IgG levels determined with this assay correlated with SARS-CoV-2 neutralization titers and the severity of COVID-19.

In our previous evaluation of the former Siemens COV2G test, we compared the performance characteristics of the Roche Elecsys Anti-SARS-CoV-2, Abbott SARS-CoV-2 IgG, Siemens SARS-CoV-2 total (COV2T) and SARS-CoV-2 IgG (COV2G), and found a markedly lower sensitivity of the Siemens COV2G (78.8%) compared to all other assays. In contrast, the new Siemens sCOVG was significantly more sensitive (90.5%). This is well in line with the sensitivities observed for all other assays evaluated with our cohort (range 90.4% to 93%) [19]. Additionally, the sensitivity is essentially in line with the manufacturer’s claims, which state a sensitivity of 91.14% for samples between day 14-20 after PCR-diagnosis and 96.41% for samples dated ≥ 21 days after PCR diagnosis (Supplemental Table S1). Still, the rate of COVID-19 patients without detectable antibody response with various assays in our cohort is higher than reported by several manufacturers. On the one hand, this might be partly explained by the inclusion of patients with immunosuppression in our study [19]. In this regard, 9 out of the 13 samples which gave concordantly negative results with four diverse serologic assays in our previous study (mainly due to immunosuppressive therapy of those patients, including chemotherapy, anti-CD20 antibodies and cortisone), were included in this study and all resulted negative with the sCOVG assay too. On the other hand, a number of manufacturer independent studies reported real-life data that are well comparable to our results [24-29]. Specificity was >99% and thus within the specifications of the manufacturer. We observed no obvious differences in specificities between the COV2G and the sCOVG test although our study was not powered to detect small differences. Thus, the performance characteristics regarding specificity and sensitivity of the Siemens sCOVG assay are basically in line with those observed for other fully automated chemiluminescence immunoassays run on high throughput random access analysers [15-20] and with the information given by the manufacturer.

We previously found a lower rate of antibody positivity in the COV2G in females than in males [19]. This difference was not significant for the new sCOVG assay as the clinical sensitivity was improved for all sub-cohorts. In this regard, the sCOVG seems to detect the previous COVID-19 infection more robustly. However, the quantitative S1-RBD IgG levels were significantly higher in males compared to females in our study, which is in line with higher anti-S- and N-antibodies [30] or higher anti-S-antibodies and neutralizing antibodies in male than in female subjects [31] in other studies, respectively. Similarly, a significantly lower rate of antibody positivity has been observed for outpatients compared to hospitalized patients with the former COV2G assay [19] but not with the new sCOVG assay. Again, the quantitative S-RBD IgG levels correlated with the severity of the disease in our study. Rijkers et al. found higher RBD total antibodies and higher neutralizing antibody titers in severe (hospitalized) vs. mild (non-hospitalized) COVID-19 patients [32]. Also, other authors described that severe COVID-19 patients had a more vigorous IgG [33] and higher neutralizing antibody response [34]. This would be in line with the observation of higher levels of antibodies in elderly patients as they have a higher prevalence of complicated disease [23]. Moreover, complicated disease is associated with more pronounced immune activation and sustained inflammation [35, 36], which may translate into more sustained immune responses and higher antibody titers. However, some authors did not find an association between antibody response [37] or neutralizing antibody response [38] and disease severity. The reasons for the discrepancies between these reports remain unknown but may involve differences in the study design, patient cohorts or types of immunoassays used.

The quantification of SARS-CoV-2 IgG levels is an additional benefit of the sCOVG assay compared to its predecessor COV2G and may aid to monitor the antibody levels after COVID-19 or after vaccination over time [39-42]. In this regard, further harmonization of antibody measurement is ongoing to standardize the monitoring of humoral immune response in the future and to estimate the degree of protection and to predict its likely duration [18]. Assays detecting antibodies against the RBD of the S-Protein of SARS-CoV-2 might be of particular interest as they also detect neutralizing antibodies interfering with the binding of the SARS-CoV-2 to the ACE receptor [14, 15]. In our study, we found a good qualitative and quantitative correlation of the sCOVG result with a SARS-CoV-2 virus neutralization assay in 134 COVID-19 patients. Only two (1.5%) out of all 134 samples showed discrepant qualitative results, and in both cases the neutralization assay was positive while the sCOVG remained below the threshold for positivity. Conversely, all sCOVG-positive samples had a positive neutralization assay result. However, the pre-test probability needs to be considered, as this part of our study was limited to patients recovered from COVID-19 and who thus had a high likelihood of having mounted a neutralizing antibody response. In other cohorts including individuals without COVID-19 infection or vaccination, the potential of false positive results needs to be considered for serologic testing. SARS-CoV-2 IgG has been shown to correlate with virus neutralizing titers [43]. Moreover, S-protein based immunoassays correlate better with neutralizing activity than N-protein based assays [15]. For the S1-RBD based Siemens sCOVG, we found a higher correlation with a neutralization assay than other authors for other immunoassays [15-17].

It is currently unclear if serological testing is of clinically need for individuals after COVID-19 vaccination. IgG against the S-protein of SARS-CoV-2 is typically found after vaccination [44-46]. Prerequisites for assays for estimating the humoral immune response to COVID-19 vaccinations at the individual level include the usage of the correct antigen (e.g. the S-protein for the mRNA-based vaccines BNT162b2 from Biontech/Pfizer and mRNA-1273 from Moderna and for AstraZenecas adenoviral vector-badsed vaccine ChAdOx1 [40-42]) and the correct antibody isotype (IgG, due to their longevity [47]), the potential to quantify the results and a good correlation of antibody results with the presence of neutralizing antibodies. The sCOVG test potentially fulfills all those criteria.

Our preliminary experience in BNT162b2 (BioNTech/Pfizer) vaccine recipients suggests, that the Siemens sCOVG assay may be suitable to monitor the humoral immune response to vaccination with this vaccine (data not shown). Reactivity in the sCOVG is seen already two to three weeks after the first vaccine dose in individuals previously not infected with SARS-CoV-2 and very high sCOVG values are seen one week after the first dose in recovered COVID-19 patients. However, further studies with standardized quantification of SARS-CoV-2 S-RBD IgG are expected to provide a useful surrogate for virus neutralization capacity and to establish the basis for clinically relevant antibody level cut-offs after SARS-CoV-2 infection and vaccination. Ideally, the peak levels and dynamics of antibody levels will enable us to predict the extent and the duration of immunity against COVID-19.

In summary, we performed an independent clinical evaluation of the quantitative Siemens SARS-CoV-2 IgG (sCOVG) assay. To the best of our knowledge, this is the first published external validation of this test for IgG antibodies against the S-RBD of SARS-CoV-2. The assay showed improved sensitivity compared to the predecessor test COV2G. Overall, specificity and sensitivity of the sCOVG assay are comparable to those observed for other fully automated chemiluminescence immunoassay tests on high throughput random access analysers in our cohort. Comparisons with the results from a SARS-CoV-2 neutralization assay indicate a good correlation with the sCOVG S1-RBD IgG levels in convalescent COVID-19 patients. In the future, the quantification of SARS-CoV-2 S-RBD IgG antibody response will be of interest not only to monitor the humoral immune response after COVID-19 disease but also upon vaccination.

## Data Availability

Data available on request from the authors

## Abbreviations

CI: confidence interval
COI: cut-off index
COVID-19: Coronavirus disease 2019
CV: coefficient of variation
ICU: intensive care unit
Ig: immunoglobulin
N: nucleocapsid protein of the SARS-CoV-2 virus
RBD: receptor binding domain
RT-PCR: reverse transcription polymerase chain reaction
S: spike protein of the SARS-CoV-2 virus
SARS-CoV-2: Severe Acute Respiratory Syndrome Coronavirus 2
TCID_50_: Tissue Culture Infection Dose 50 (median tissue culture infectious dose)
WHO: World Health Organization

## Acknowledgements

The authors thank Gernot Osterer (Siemens) for technical support.

## Research funding

The study was performed by institutional research funding.

## Author contributions

CI, AE, WP, LL, CJB, MA, AG, and GH performed or analysed serological tests. BHo and BHa performed virus neutralization tests. SS, AP, TS, WM, HS, JL-R, RB-W, IT, and GW obtained clinical data. CI and AE performed statistical analysis. CI, AE, AG, and GH designed the study and wrote the paper. All authors revised and approved the manuscript.

## Financial

IT was awarded an Investigator-Initiated Study (IIS) grant by Boehringer Ingelheim (IIS 1199-0424).

