## Supplemental material for "Clinical validation of the quantitative Siemens SARS-CoV-2 spike IgG assay (sCOVG) reveals improved sensitivity and a good correlation with virus neutralization titers"

### Supplemental Tables

**Supplemental Table S1.** Characteristics of 134 COVID-19 patients (sCVOG - neutralisation assay correlation cohort)

| | | Absolute | % |
|---|---|---|---|
| **Number** | n | 134 | 100% |
| **Disease severity** | Outpatient | 26 | 19% |
| | General ward | 68 | 51% |
| | ICU | 31 | 23% |
| | Not known | 9 | 7% |
| **Age (years)** | min | 20 | |
| | max | 88 | |
| | median (IQR) | 57 (50- 68) | |
| **Sex** | female | 58 | 43% |
| | male | 76 | 57% |
| **Symptom onset known** | yes | 120 | 90% |
| | no | 14 | 10% |
| **Date of first positive PCR known** | yes | 122 | 91% |
| | no | 12 | 9% |
| **Time between symptom onset and PCR (days)**[a] | median (IQR) | 5 (2-8) | |
| **Time between disease onset and blood draw (days)**[b] | median (IQR) | 63 (54-74) | |

[a] Refers to the first positive SARS-CoV-2 PCR of the patient; [b] refers to the representative sample used in the sensitivity analysis. Abbreviations: IQR, interquartile range, PCR, polymerase chain reaction.

**Supplemental Table S2.** Attributes for the two Siemens SARS-CoV-2 IgG assays as given by the manufacturer's package inserts.

| Attribute | SARS-CoV-2 IgG (sCOVG) | SARS-CoV-2 IgG (COV2G) |
|---|---|---|
| **Manufacturer** | **Siemens** | **Siemens** |
| **Version of the package insert** | October 2020 | June 2020 |
| **CE-IVD status** | Conform | Conform |
| **EUA status** | Granted | Granted |
| **Method** | CLIA | CLIA |
| **Platform** | ADVIA Centaur XP [a], XPT | ADVIA Centaur XP [a], XPT |
| **Specimen type** | Serum, plasma (Li-heparin), venipuncture or capillary puncture | Serum, plasma (Li-heparin, K-EDTA) |
| **Sample volume (without dead volume)** | 40 µl | 10 µl |
| **Detected antibodies** | IgG | IgG |
| **Solid-phase antigen target** | RBD of the S1 protein | RBD of the S1 protein |
| **Result calculation** | Index | Index |
| **Interpretation** | <1,0: negative<br>≥1,0: positive<br>1.00 Index value = 1.00 IU/ml | <1,0: negative<br>≥1,0: positive |
| **Diagnostic specificity*** | 99.90% (99.64-99.99, n=1,995) | 99.89% (99.61-99.99, n=1,831) |
| **Diagnostic sensitivity*** | *Days after first PCR-positivity*<br>0-6: 50.82% (45.58-56.03, n=368)<br>7-13: 82.47% (76.38-87.55, n=194)<br>14-20: 91.14% (82.59-96.36, n=79)<br>≥21: 96.41% (92.74-98.54%, n=195) | *Days after first PCR-positivity*<br>0-6: 53.49% (42.41-64.23, n=86)<br>7-13: 93.44% (84.05-98.18, n=61)<br>≥14:100.00% (91.59-100.00%). n=42 |
| **Accuracy** | Within-day: 2.6-5.8%<br>Between-day: 3.9-6.7% | Within-day: 2.0-4.6%<br>Between-day: 2.8-6.1% |

In brackets 95% confidence intervals are shown. Abbreviations: CE-IVD, European Conformity - In Vitro Diagnostic Medical Device; CLIA, chemiluminescence immunoassay; EUA: Emergency Use Authorization; K, potassium; Li, Lithium; n: number of samples tested; RBD: receptor-binding domain; S1: Spike 1 protein of the SARS-CoV-2 virus; [a] indicates the platform used in our study.

**Supplemental Table S3.** Measurements of the immunoglobulin formulation Privigen®, undiluted and 1:50 diluted.

| Privigen | Siemens SARS-CoV-2 IgG (sCOVG) | Siemens SARS-CoV-2 IgG (COV2G) |
|---|---|---|
| Undiluted | 1.28 | 1.09 |
| 1:50, negative serum | 0.00 | 0.01 |
| 1:50, sodium chloride | 0.00 | 0.23 |

Data indicate the respective results of the raw data indexes of the tests. Privigen® was diluted with negative pre-pandemic serum or sodium chloride, respectively.

**Supplemental Table S4.** Diagnostic sensitivity of the two assays stratified for disease course, age and gender.

| | | Siemens SARS-CoV-2 IgG (sCOVG) | Siemens SARS-CoV-2 IgG (COV2G) |
|---|---|---|---|
| **Sensitivity according to PI**[a] | | **91.1% (82.6-96.4%)** | **100.00% (91.59-100.00%)** |
| **All patients** | Total | 178 | 178 |
| | Positive | 161 | 140 |
| | Negative | 17 | 38 |
| | **Sensitivity** | **90.5% (85.2-94.3)** | **78.7% (71.9-84.4)** |
| **Male** | Total | 113 | 113 |
| | Positive | 105 | 98 |
| | Negative | 8 | 15 |
| | Sensitivity | 92.9% (86.5-96.9) | 86.7% (79.1-92.4) |
| **Female** | Total | 65 | 65 |
| | Positive | 56 | 42 |
| | Negative | 9 | 23 |
| | Sensitivity | 86.2% (75.3-93.5) | 64.6% (51.8-76.1) |
| **18-49** | Total | 59 | 59 |
| | Positive | 51 | 38 |
| | Negative | 8 | 21 |
| | Sensitivity | 86.4% (75.0-94.0) | 64.4% (50.9-76.5) |
| **50-69** | Total | 82 | 82 |
| | Positive | 76 | 71 |
| | Negative | 6 | 11 |
| | Sensitivity | 92.7% (84.8-97.3) | 86.6% (77.3-93.1) |
| **70-100** | Total | 37 | 37 |
| | Positive | 34 | 31 |
| | Negative | 3 | 6 |
| | Sensitivity | 91.9% (78.1-98.3) | 83.8% (68.0-93.8) |
| **Outpatient** | Total | 65 | 65 |
| | Positive | 55 | 41 |
| | Negative | 10 | 24 |
| | Sensitivity | 84.6% (73.5-92.4) | 63.1% (50.2-74.7) |
| **Gen. ward** | Total | 85 | 85 |
| | Positive | 79 | 72 |
| | Negative | 6 | 13 |
| | Sensitivity | 92.9% (85.3-97.4) | 84.7% (75.3-91.6) |
| **ICU** | Total | 28 | 28 |
| | Positive | 27 | 27 |
| | Negative | 1 | 1 |
| | Sensitivity | 96.4% (81.7-99.9) | 96.4% (81.7-99.9) |

Sensitivity of the evaluated test-systems including all SARS-CoV-2 PCR-confirmed patients (one sample per patient; the sample ≥14 days and closest to day 28 after symptom onset was used) and stratified for severity of the disease (outpatient, hospitalised at the general ward, hospitalized at the intensive care unit) as well as age (18-49, 50-69, 70-100 years), respectively. 95% confidence intervals are given in brackets. Abbreviations: PI: package insert. [a] ≥ day 14 after PCR-diagnosis.

**Supplemental Table S5.** Qualitative agreement between the Siemens SARS-CoV-2 IgG and the neutralisation assay.

| | **Neutralisation assay** | |
|---|---|---|
| **sCOVG** | negative | positive |
| negative | 8 | 2 |
| positive | 0 | 124 |

### Supplemental Figures

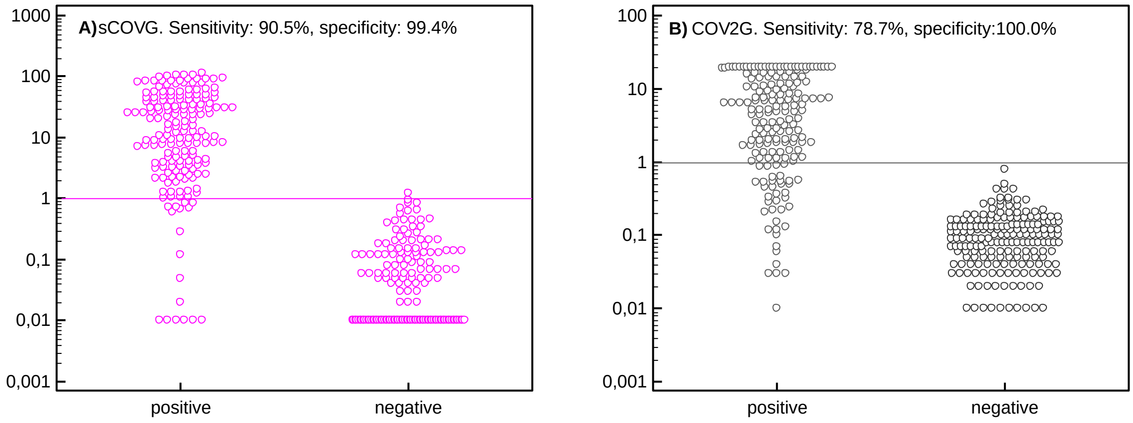


**Supplemental figure S1.** Raw data indexes for the investigated tests Siemens SARS-CoV-2 IgG sCOVG (A) and COV2G (B) of the positive samples (178 samples of different SARS-CoV-2 PCR-confirmed patients) and negative samples (pre-pandemic samples, sCOVG n = 160, COV2G n = 191). The horizontal lines in the dot plots represent the manufacturer's cut off index.

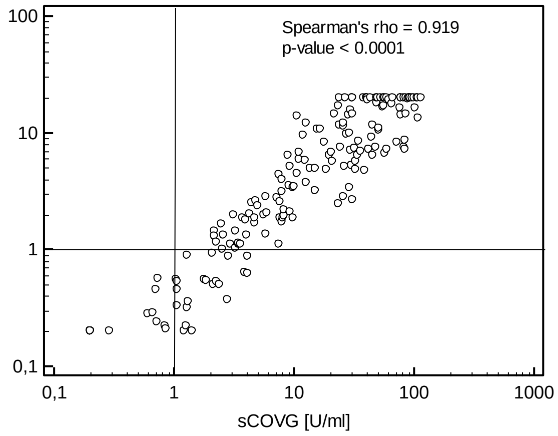


**Supplemental Figure S2.** Scatter diagram of the quantitative values of the Siemens SARS-CoV-2 IgG sCOVG and the COV2G assays. For illustration purposes, values below 0.2 U/ml or Index, respectively, were set as 0.2. Take note: the upper measurement limit of the COV2G assay is 20.0 Index.

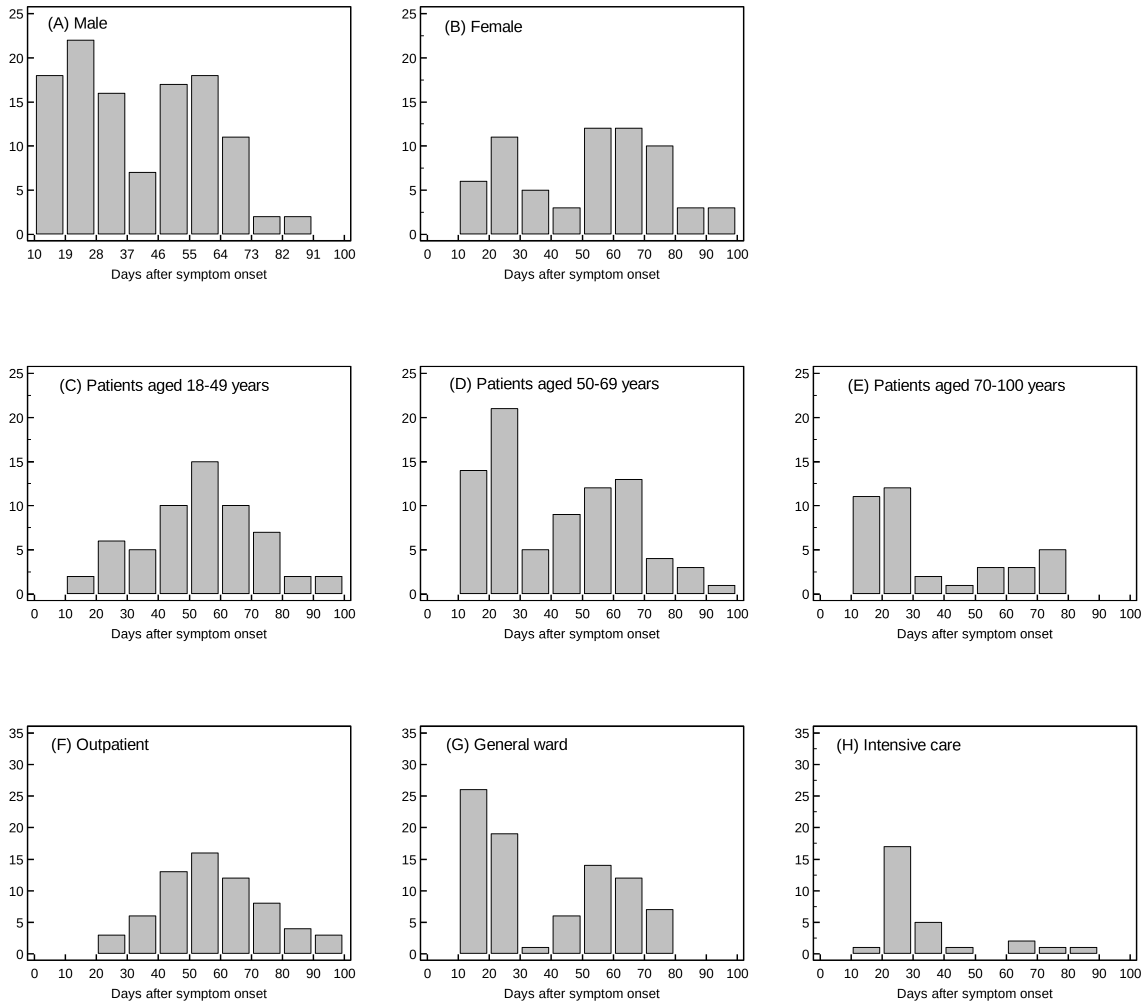


**Supplemental Figure S3.** Frequency distribution (histograms) of the sample timing used for sensitivity and quantitative results analyses in the different sub-cohorts.
